# Bone mineral density assessment in patients with cystinuria

**DOI:** 10.1101/2023.12.06.23299590

**Authors:** Viola D’Ambrosio, Giovanna Capolongo, Chiara Caletti, Maria Teresa Vietri, Martina Ambrogio, Gianmarco Lombardi, Alessandra F Perna, Giuseppe Orefice, Elisa Gremese, Valentina Varriano, Davide Gatti, Angelo Fassio, Giovambattista Capasso, Giovanni Gambaro, Pietro Manuel Ferraro

## Abstract

**Introduction:** Cystinuria is a rare genetic disease characterized by impaired tubular transport of cystine. Clinical features of cystinuria include mainly nephrolithiasis and its complications, although cystinuric patients may present with other comorbidities. There are currently no data on bone features of patients with cystinuria. Our aim is to characterize bone mineral density (BMD) in cystinuria.

**Methods:** Our study included adult cystinuric patients followed at 3 specialized outpatient clinics in Italy (Rome, Naples and Verona). Markers of bone turnover were analyzed in a centralized laboratory. Clinical, biochemical and dual-energy X-ray absorptiometry (DEXA) data were collected from September 2021 to December 2022. One sample t-tests were used to assess whether BMD Z-scores were significantly different from 0.

**Results:** 39 patients were included in the study. Mean (SD) age was 38 (15) years, 40% were women. Mean estimated glomerular filtration rate was 92 mL/min/1.73 m^2^. Serum parameters associated with bone remodeling (parathyroid hormone, FGF23, calcium and phosphate) were all in the normal range, with only 4 patients showing mild hypophosphatemia. Prevalence of low BMD, defined as T-score < −1 at any site, was 65%, higher at the femur and lumbar spine. Average Z-scores were negative across most sites.

**Conclusions:** Our data show a high prevalence of low BMD in cystinuric patients, despite having normal (or moderately impaired) kidney function and being relatively young.

## Introduction

Cystinuria is a rare genetic disease characterized by impaired tubular transport of cystine and other dibasic amino acids^1^. Despite being a rare condition, with an average prevalence of 1 in 7,000 live births, cystinuria is considered the most frequent monogenic cause of nephrolithiasis, accounting for 1-2% of kidney stones in adults and about 8% in children^2^.

Cystine is a homodimer of the amino acid cysteine that is absorbed in the gastrointestinal tract, freely filtered by the glomerulus and then reabsorbed in the proximal tubule. In patients affected by cystinuria, a defective apical transporter located in the proximal tubule leads to an increased urinary excretion of cystine and therefore to formation of cystine calculi. Besides cystine, other amino acids such as ornithine, lysine and arginine are not properly reabsorbed by the proximal tubule, but their solubility is higher than that of cystine and they do not lead to nephrolithiasis.

Cystinuria is currently classified as type A and type B based on the gene involved. SLC3A1 encodes for the heavy subunit rBAT of the rBAT-b0,+AT amino acid transporter system in intestinal and proximal tubule cells, and is responsible for cystinuria type A. SLC7A9 encodes for the light subunit of the rBAT-b0,+AT, and it accounts for cystinuria type B^3^. Rare cases of patients with mutations in both genes have been reported (type AB). Heritability of the disease is autosomal recessive.

Cystinuric patients have a high prevalence of chronic kidney disease (CKD) and kidney failure, not only compared to the general population, but also compared to other stone formers^2^. This is partially explained by recurrent nephrolithiasis, urinary tract and Bellini’s ducts obstruction, recurrent urinary tract infection and hypertension; the latter is another common finding in cystinuric patients^4^. Being a consequence of CKD^5(p1)^, decreased bone mineral density (BMD) can potentially affect cystinuric patients. However, its frequency and whether this is attributable to secondary hyperparathyroidism as a consequence of loss of glomerular filtration rate (GFR) alone is still uncertain, and more evidence on humans is needed. In a study conducted by Peters et al. in 2003 on a murine model, SLC3A1 knock-out mice were characterized by reduced BMD irrespective of the presence or absence of uremia, suggesting a possible bone metabolic alteration at a very early stage in cystinuria even in the absence of overt kidney damage^6^. A strong correlation between stone formation and elevated alkaline phosphatase was also noticed, suggesting a high bone turnover in this condition^6^. Our project aims at investigating bone abnormalities in a cohort of patients with cystinuria.

## Methods

39 adult patients were enrolled: 14 from Fondazione Policlinico Universitario A. Gemelli IRCCS in Rome, 13 from Azienda Ospedaliera Universitaria, Università degli Studi della Campania in Naples, and 12 from Azienda Ospedaliera Universitaria Integrata in Verona. Blood was drawn from each patient in one serum and one EDTA-plasma tube, immediately centrifuged at 4500 rpm for 15 minutes and frozen locally at −80°C. The batch was then shipped to a centralized laboratory in Naples where markers of bone metabolism (calcium, phosphate, parathyroid hormone [PTH], fibroblast growth factor 23 [FGF23])^7^ and serum creatinine were measured. Additional electronic records for serum biochemistry, urinary parameters and DEXA scans were retrospectively collected and analyzed. Body mass index (BMI) was calculated with the formula weight (in kg)/square of height (in m) and was considered to be normal between 18.5 and 24.9 kg/m^2^. Estimated GFR (eGFR) was calculated with the 2009 CKD-EPI formula and was considered to be normal when ≥60 mL/min/1.73 m^2^. Hypertension was defined as systolic blood pressure (SBP) ≥ 140 mmHg or diastolic blood pressure (DBP) ≥ 90 mmHg, according to the most recent National Institute for Health and Care Excellence (NICE) guidelines or based on antihypertensive treatment. Patients were defined as osteopenic or osteoporotic according to the World Health Organization (WHO) definition of T-score between −1 and −2.5 SD and T-score < −2.5 SD at any site, respectively. Low BMD was defined as a T-score < −1. All participating laboratories and imaging centers comply with current international image management and quality controls rules and regulations. In order to evaluate statistically significant departures from zero of T-scores and Z-scores (which can be interpreted as significant differences compared with a reference and an age- and sex-matched population, respectively), we used linear regression models with random effects for center to incorporate systematic clustering of patients. A two-tailed p-value <0.05 was considered statistically significant. The study has Ethical Committee approval (Prot. 0006896/e, 26/03/2020 Comitato Etico Università degli Studi della Campania “Luigi Vanvitelli” - Azienda Ospedaliera Universitaria “Luigi Vanvitelli”-AORN “Ospedali dei Colli”; Prot. 3229 11/03/2021 Comitato Etico Fondazione Policlinico Gemelli; Prot n. 38525 28/06/2021 Comitato Etico Università degli Studi di Verona). Adult (≥18 years) patients affected by cystinuria with eGFR ≥15 mL/min/1.73 m^2^ who signed the informed consent were included. Patients who did not sign informed consent, pregnant women, patients undergoing chronic dialysis and patients with previous renal transplant were all excluded. The study has been conducted in accordance to the declaration of Helsinki.

## Results

Descriptive data are reported in Table 1. 40% of patients were women, of whom about one third was in menopause. Our cohort had a mean (SD) age of 38 (15) years, with a BMI of 25 (6) kg/m^2^ and eGFR of 92 (22) mL/min/1.73 m^2^. Only 3 patients had a previous diagnosis of osteoporosis, whereas 7 patients reported a history of previous fracture, all of them traumatic. CKD, defined as eGFR <60 mL/min/1.73 m^2^, had a prevalence of 8% whereas hypertension 17%. One patient had undergone previous left nephrectomy for staghorn calculi.

**Table 1.** Baseline characteristics of the study cohort. Continuous variables are reported as means and standard deviations and categorical variables as frequencies and percentages. *BMI* body mass index; *eGFR* estimated glomerular filtration rate, *CKD* chronic kidney disease.

| Characteristics | Means and Frequencies |
| --- | --- |
| Age (years) | 38 (15) |
| BMI (kg/m<sup>2</sup>) | 25 (6) |
| eGFR (mL/min/1.73 m<sup>2</sup>) | 92 (22) |
| Women | 15, 40% |
| Menopause (n, % of women) | 5, 33% |
| Previous diagnosis of osteoporosis | 3, 8% |
| Previous fractures | 7, 18% |
| Traumatic (n, % of fractures) | 7, 100% |
| CKD (eGFR <60 mL/min/1.73 m<sup>2</sup>) | 3, 8% |
| Hypertension | 17% |

Data regarding relevant therapy to both nephrolithiasis and bone health are reported in Table 2. Cholecalciferol was the treatment of choice in all people treated with oral vitamin D (23%, average dose 26,000 IU/month). Treatment with tiopronin (average dose 980 mg/day) was less frequent than oral potassium citrate supplements (average dose 49 mEq/day). Nine patients were prescribed additional oral alkali treatment.

**Table 2.** Frequency of relevant treatments in the study cohort.

| Therapy | Frequency |
| --- | --- |
| Cholecalciferol | 9, 23% |
| Bisphosphonate | 0 |
| Tiopronin | 15, 38% |
| Potassium citrate | 34, 87% |
| Additional alkali therapy | 9, 23% |
| Sodium bicarbonate | 7, 78% |
| Calcium carbonate | 2, 22% |

Overall, patients had relatively preserved kidney function with normal eGFR. 3 patients (8%) had an eGFR <60 mL/min/1.73 m^2^. Patients had also serum PTH, calcium, phosphate and FGF23 values generally in the normal range, except for 2 patients with low FGF23 and 4 patients with hypophosphatemia (Figure 1 and Supplementary Table 1).

**Figure 1.**
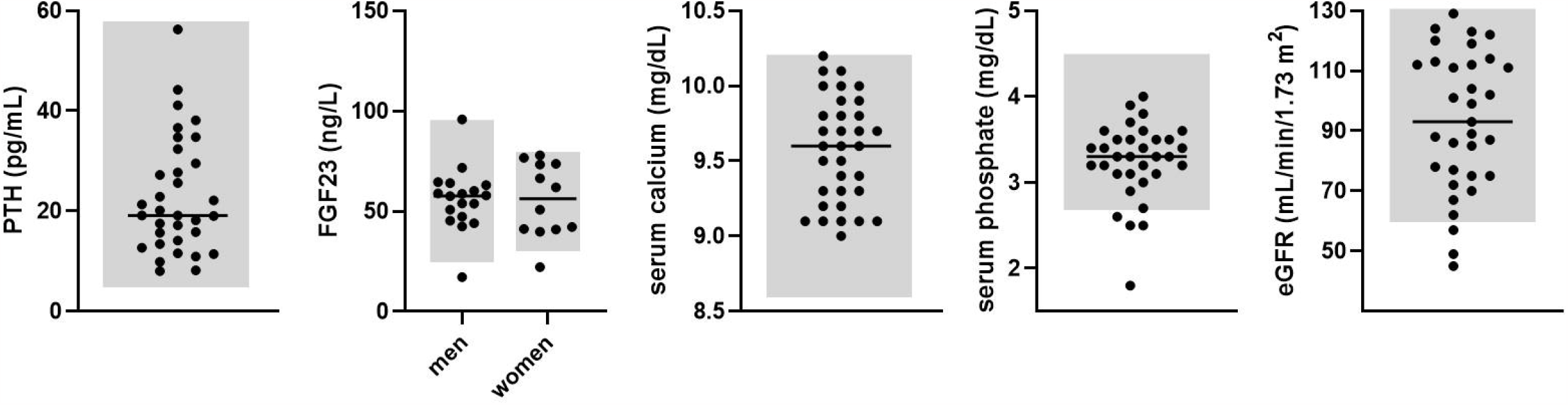
Dot plots of serum biomarkers measured in the study cohort; the grey areas represent normal ranges; the black lines represent median values. *PTH* parathyroid hormone; *FGF23* fibroblast growth factor 23; *eGFR* estimated glomerular filtration rate.

An additional analysis was then performed on urinary metabolic assessment and additional serum biochemistry of our cohort (Supplementary Table 2).

DEXA data were available for 30 patients. Prevalences of osteopenia and osteoporosis are shown in Table 3; overall, the prevalence of low BMD was 65% (50% and 15% with osteopenia and osteoporosis, respectively). Higher prevalence of osteopenia was found in femoral ward triangle (39%), followed by L1 (37%), L2 and L4 (both 33%). Osteoporosis was mainly prevalent at the femur’s neck and ward triangle.

**Table 3.** Prevalence of osteopenia and osteoporosis in the study cohort.

| Site | Prevalence of osteopenia | Prevalence of osteoporosis |
| --- | --- | --- |
| L1 | 37% | 3% |
| L2 | 33% | 0% |
| L3 | 11% | 3% |
| L4 | 33% | 0% |
| L1-L4 | 22% | 0% |
| L2-L4 | 18% | 0% |
| Femoral | 27% | 0% |
| Neck | 20% | 12% |
| Ward | 39% | 13% |
| Trochanter | 21% | 0% |
| Any | 50% | 15% |

Average T-score values across all sites were negative except for L3 and L4. Average Z-score values were negative in L1, L1-L4, L2-L4, femoral neck, ward and trochanter. To test whether T- and Z-scores were significantly negative and different from 0, linear regression analysis was performed across every site. This difference was confirmed to be statistically significant for L1, femoral neck and ward T-scores mean values. Across Z-scores, no site reached statistical significance. Average Z-score values and 95% confidence intervals are summarized in Figure 2. Prevalence of osteopenia and osteoporosis stratified by age are shown in Table 4.

**Table 4.** Prevalence of osteopenia and osteoporosis stratified by age groups.

| Age (years) | n of patients for age range | Prevalence of osteopenia | Prevalence of osteoporosis |
| --- | --- | --- | --- |
| 20-39 | 16 | 50% | 0% |
| 40-59 | 7 | 57% | 14% |
| ≥60 | 5 | 60% | 40% |
| Overall | 28 | 50% | 15% |

**Figure 2.**
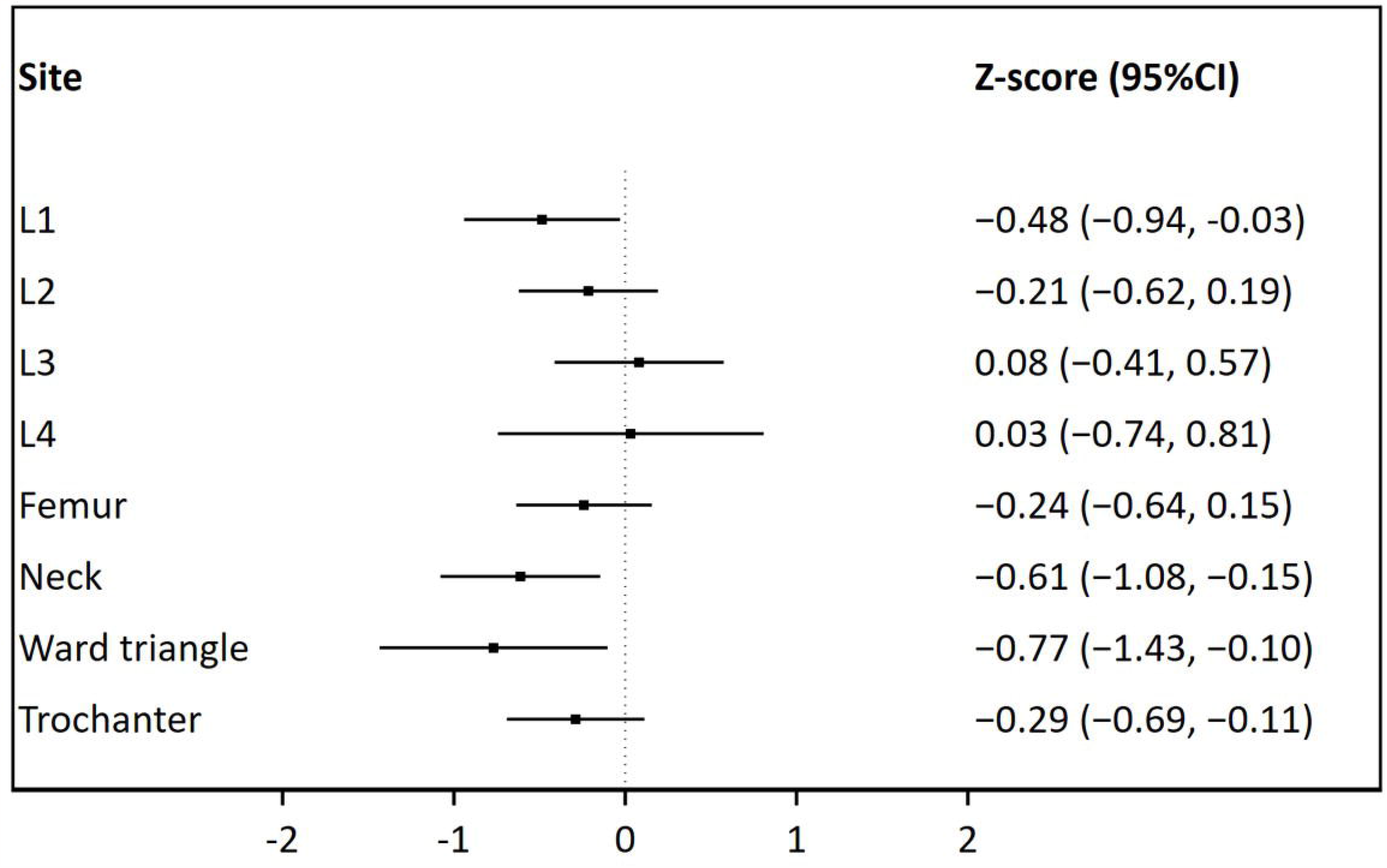
Forest plot showing average Z-score values with 95% confidence intervals.

## Discussion

Our study shows that cystinuric patients have a high prevalence of low BMD, especially when stratified by age group. Although these are just descriptive statistics, evidence from other cohorts in the literature provides help in critically analyzing these data. The study conducted by Bijelic R et al^8^ is particularly interesting in this regard and allows us to highlight the remarkably high prevalence of low BMD in cystinuric patients compared to stone formers and controls (Supplementary Table 3). The relationship between nephrolithiasis and low bone mineral density (BMD) has been extensively studied. These considerations have shaped the way we look at nephrolithiasis, from a strictly urological, to a metabolic and systemic condition. Low BMD is indeed associated with increased risk of fractures^9^, morbidity and mortality, that worsen kidney stone formers’ prognosis^10,11^. Previous meta-analyses have confirmed the association between low BMD and nephrolithiasis^12,13^, especially in the presence of hypercalciuria. This is not surprising given the well-known link between negative calcium balance and reduced BMD. Interestingly, increased excretion of urinary calcium had a low prevalence in our cohort, suggesting that negative calcium balance is not the main driver for low BMD in cystinuria. In addition, no overt altered serum bone remodeling parameters were found, although the prevalence of fractures suggests a predisposition to bone frailty, considering that our cohort is rather young and with preserved kidney function. It should also be noted that a high percentage of patients were treated with potassium citrate (a smaller one with additional alkali therapy) (Table 2), known to have a positive effect on bone mineralization^14,15^. We could therefore speculate that BMD would be even lower in the absence of oral alkali supplementation. Prevalence of CKD and hypertension were found to be much lower compared to data in the literature ^2,16,17,4^. Although, this is probably due to the high standard of care these patients are provided with in highly specialized centers. It is therefore likely that there might be other underlying mechanisms explaining loss of BMD in cystinuric patients. We hypothesized mechanisms that could explain reduced bone mineral density in cystinuric subjects. The first one is based on cystinuria pathophysiology. Cystinuric patients have increased excretion of three dibasic amino acids of which lysine is an essential amino acid, arginine is an essential amino acid under certain condition (infancy and adolescence) and ornithine is a derivate of arginine. Taken together, these three amino acids are precursor or intermediate metabolites of the urea cycle in hepatocytes, a pathway that is essential for waste nitrogen products’ formation and therefore disposal. rBAT/b^+,0^ is expressed also in the gut and previous experiments demonstrated that cystinuria is characterized by a defective intestinal amino acids transport^18,19^. Taken together, this causes low plasma arginine that is thought to be compensated by an increase in alanine and glutamate^20,21,22^, two precursor amino acids of Krebs cycle^23^. We hypothesize that the reduced availability of arginine in the urea cycle causes an activation of Krebs cycle via the arginine-succinate shunt and the release of alanine and glutamate from muscle. At steady state, muscle catabolism may be minimal but could account for sarcopenia predisposition in cystinuric patients. Ultimately, sarcopenia is a known risk factor for osteopenia ^24^. The second hypothesis is strongly related to the first one and it involves citrate. Citrate is an intermediate metabolite of Krebs cycle which is essential for cell survival^25^. The previously-hypothesized increase in Krebs cycle’s activation, to compensate for urea cycle intermediates’ loss in cystinuria, could therefore lead to increased release of citrate from the bone where it stabilizes apatite crystals^26^. This could ultimately predispose to bone demineralization. The third hypothesis involves acid-base balance and dietary acid load. In a urine proteomic profiling study conducted in patients with cystinuria, it was demonstrated that ammonium transporters are down-regulated^27^. This suggests that the physiological response of the kidney to an increased potential renal acid load (PRAL) may be altered. Dietary habits could also play a role not only in the worsening of cystinuria itself, but also in the progression of CKD and stone formation. Additional evidence comes from Sakhaee K et al ^28^ who performed an acid load test (NH_4_Cl) on 5 cystinuric patients that were hypocitraturic and had a urinary pH >6.5. These patients had indeed a urinary acidification defect.

Among the strengths, our cohort is a rather large cohort for a rare condition and includes 3 of the biggest stone centers in Italy and represents the first evidence of bone disorders in cystinuria. Our study has also some limitations. First of all, it lacks a control cohort.

DEXA results are however expressed as standard deviations from a comparator (for T-score: bone density of a healthy 30-year-old person; for Z-score: average bone density of people of the same age, sex and BMI). Another limitation is the fact that DEXA scans were performed in different centers and might be operator-dependent. Urinary and serum biochemistry were retrospectively collected and were not performed in the same laboratory. Among biochemical parameters, serum bicarbonate and alkaline phosphatase were not available, although it would have been interesting to investigate them.

## Conclusions

To our knowledge this is the first study to systematically evaluate mineral bone density in cystinuric patients. Our findings suggest that there is a tendency towards decreased bone mineral density irrespective of the presence of low eGFR. The underlying mechanisms that could explain our findings have not been directly investigated, although we speculate that metabolic abnormalities and concomitant defective tubular transport may play a role. Our findings suggest a shift in the way we consider cystinuria, from a kidney-specific disease to a systemic and metabolic condition. We therefore suggest a complete metabolic work-up in patients with cystinuria, including bone health serum parameters even in the absence of CKD to monitor for early signals of bone disorders.

## Data Availability

Data will be available upon reasonable request to the corresponding author.

## Acknowledgments

We are grateful to the patients and their families for the constant support in research

## Data Availability Statement

Data will be available upon reasonable request to the corresponding author.

## Supplementary material

Supplementary Table 1. Descriptive statistics of serum biomarkers measured in the study cohort. PTH parathyroid hormone; FGF23 fibroblast growth factor 23; eGFR estimated glomerular filtration rate.

Supplementary Table 2. Descriptive statistics for urinary and serum biochemistry of our cohort. *Collected data were not from the same laboratory therefore a unique normal range could not be defined.

Supplementary Table 3. Prevalence of low BMD stratified per age group in our cystinuric cohort, in stone formers and controls from the study conducted by Bijelic R et al6. BMD bone mineral density.

