## Supplementary material for "Bone mineral density assessment in patients with cystinuria"

| Biomarker | Mean | Standard deviation | Median | 25° percentile | 75° percentile | Missing data | Normal values |
|---|---|---|---|---|---|---|---|
| **PTH (pg/mL)** | 22.9 | 11.6 | 19.1 | 13.7 | 30.9 | 6 | 4.6-58.1 |
| **FGF23 (ng/L)** | | | | | | | |
| **Men** | 55.9 | 15.5 | 57.7 | 46.8 | 63.2 | 6 | 25.1-95.5 |
| **Women** | 55.5 | 18.4 | 56.3 | 41.0 | 73.5 | 3 | 30.8-79.6 |
| **Calcium (mg/dL)** | 9.57 | 0.35 | 9.60 | 9.25 | 9.85 | 6 | 8.6-10.2 |
| **Phosphate (mg/dL)** | 3.24 | 0.44 | 3.30 | 3.10 | 3.50 | 6 | 2.7-4.5 |
| **eGFR (mL/min/1.73 m$^2$)** | 92 | 22 | 93 | 75 | 111 | 6 | >60 |

**Supplementary Table 1**. Descriptive statistics of serum biomarkers measured in the study cohort. *PTH* parathyroid hormone; *FGF23* fibroblast growth factor 23; *eGFR* estimated glomerular filtration rate.

| Parameter | Mean | Standard deviation | Median | 25° percentile | 75° percentile | Missing data | Normal values |
|---|---|---|---|---|---|---|---|
| **Urinary cystine** mmol/24 h | 2.6 | 1.3 | 2.3 | 1.8 | 3.4 | 14 | < 0.2 |
| **Spot urinary pH** | 6.8 | 0.8 | 6.8 | 6.0 | 7.3 | 1 | - |
| **Urinary volume** mL/24 h | 2870 | 783 | 2700 | 2350 | 3200 | 10 | - |
| **Urinary calcium** mg/24 h | 155 | 93 | 128 | 112 | 180 | 8 | 100-300 |
| **Urinary phosphate** mg/24 h | 786 | 352 | 700 | 580 | 972 | 6 | 400-1300 |
| **Urinary oxalate** mg/24 h | 24 | 21 | 19 | 11 | 32 | 6 | < 40 |
| **Urinary citrate** mg/24 h | 677 | 486 | 548 | 330 | 966 | 5 | 320-1260 |
| **Urinary uric acid** mg/24 h | 456 | 171 | 450 | 340 | 600 | 9 | 250-750 |
| **Urinary sodium** mEq/24 h | 152 | 47 | 145 | 111 | 191 | 4 | 50-200 |
| **Urinary potassium** mmol/24 h | 73 | 36 | 70 | 46 | 84 | 7 | 25-125 |
| **Urinary urea** g/24 h | 23 | 12 | 20 | 15 | 26 | 4 | 26-43 |
| **Urinary creatinine** g/24 h | 1.4 | 0.4 | 1.5 | 1.0 | 1.8 | 4 | F: 0.6-1.8<br>M: 0.8-2.0 |
| **Serum potassium** mEq/L | 4.5 | 0.6 | 4.3 | 4.0 | 4.8 | 8 | 3.5-5.0 |

| | | | | | | | |
|---|---|---|---|---|---|---|---|
| **Serum urea** mg/dL | 34 | 17 | 29 | 21 | 40 | 5 | 18-45 |
| **Serum sodium** mEq/L | 140 | 3.8 | 141 | 139 | 143 | 8 | 135-145 |
| **Serum uric acid** * | 5.3 | 1.6 | 5.2 | 4.5 | 6.7 | 14 | - |

**Supplementary Table 2**. Descriptive statistics for urinary and serum biochemistry of our cohort. *Collected data were not from the same laboratory therefore a unique normal range could not be defined.

| **Low BMD prevalence** | | | |
|---|---|---|---|
| **Age group** | **Stone formers** | **Controls** | **Cystinurics** |
| **20-39** | 15% | 5% | 50% |
| **40-59** | 23.5% | 25% | 70% |
| **>60** | 45% | 40% | 100% |

**Supplementary Table 3.** Prevalence of low BMD stratified per age group in our cystinuric cohort, in stone formers and controls from the study conducted by Bijelic R et al[6]. *BMD* bone mineral density.
